# Adult hospitalizations due to hepatitis A virus infection in six tertiary hospitals across Bangladesh: December 2014 – September 2017

**DOI:** 10.1101/2025.06.26.25330300

**Authors:** Banda AA Khalifa, Repon C Paul, Arifa Nazneen, Kajal C Banik, Shariful Amin Sumon, Kishor K Paul, Arifa Akram, M Salim Uzzaman, Stephen P Luby, Emily S Gurley

## Abstract

Hepatitis A virus (HAV) is spread through the fecal-oral route. The age of infection largely determines the clinical severity; children typically have asymptomatic or mild illness, but infected adults more commonly develop clinical disease. Many countries have experienced improvements in water and sanitation which could lead to increases in the burden of HAV due to increases in the age of infection. This report summarizes the contribution of HAV to hospitalizations among patients aged >14 years from acute jaundice during 2014 - 2017 across Bangladesh. Among 1,923 patients, 148 (8%) had anti-HAV IgM antibodies in their serum; 26% were co-infected with hepatitis E or B viruses, or both, including 3 of the 4 deaths among patients with HAV infection. An outbreak of HAV was detected at one hospital during 2016. HBV vaccination was introduced in 2005, but additional hepatitis vaccination strategies may be needed to further reduce hepatitis hospitalizations and deaths.

## Introduction

Hepatitis A virus (HAV) is a picornavirus, spread through the fecal-oral route.^1^ Consequently, populations with inadequate access to clean water and food are at highest risk for transmission.^1^ While the infection is generally self-limiting and asymptomatic in children, it can cause severe illness in adults, including acute liver failure, and even death, particularly among elderly and immunocompromised individuals.^2^ According to estimates from the World Health Organization, approximately 100 million people contract hepatitis A annually, leading to 1.5 million clinical cases and 15,000–30,000 deaths.^3^

Paradoxically, areas with high transmission rates of HAV tend to have a lower burden of disease compared to areas with moderate transmission. This is because when transmission is high, nearly everyone is infected during childhood when most infections are asymptomatic or mild. Once transmission decreases, more people survive into adulthood susceptible to infection, creating a higher burden of disease if infection occurs. High income countries typically have very low burden of clinical disease, due to low transmission and protection of high risk groups through vaccination.^1^ However, many lower and middle income countries have experienced improvements in water and sanitation in recent decades, and consequently, an increased burden of disease as infections during adulthood become more common.^1^

In Bangladesh, people have historically been infected with HAV early in life. A study conducted in 2005 in Sylhet, Bangladesh found that 15% of children between 1-2 years of age already had IgG antibodies against HAV, and by age 21-25 years, 100% of the study participants had IgG antibodies.^4^ Among study participants from rural areas and lower socioeconomic backgrounds - presumably with less access to clean water - antibody prevalence reached 100% at even younger ages. Since the early 2000’s, access to clean water and improved sanitation facilities has increased in Bangladesh^5^, which could lead to decreases in HAV risk during childhood. More recently, studies have shown that the burden of HAV disease in Bangladesh may be increasing. One study of patients presenting to hospitals with acute hepatitis from 2014-2015 showed that 19% of patients aged 15-60 years had IgM antibodies against HAV, suggesting that changes in transmission could be causing increases in burden of clinical disease.^6^ However, the lack of details about severity of disease or age breakdown among these patients makes the results difficult to interpret in terms of changes to disease burden.

This analysis presents results from surveillance for hospitalizations from acute hepatitis associated with HAV among adults in six tertiary hospitals in Bangladesh from 2014 - 2017, including clinical outcomes.

## Methods

From December 2014 to September 2017, physicians reviewed hospital admission records daily to identify patients aged 14 years and older who presented with acute jaundice, defined as a new onset of yellow eyes or skin within the preceding three months; details of the surveillance methods have been published elsewhere.^7^ Each patient had blood collected and tested for serum bilirubin and serum glutamic-pyruvic transaminase levels, as well as IgM antibodies against HAV, hepatitis E virus, and surface antigens for hepatitis B virus. Information was collected from patients about their signs and symptoms and they were contacted three months post discharge to ascertain disease outcomes.

We described patients with HAV infections in terms of sociodemographics, geography, and clinical presentation, and identified risk factors for infection.

### Ethical Considerations

Written informed consent for participation was provided by patients or from their guardians if patients were seriously ill or they were aged <17 years. Patients aged 14 to 17, also provided assent for participation. The study received ethical approval from the institutional review board of the International Centre for Diarrhoeal Disease Research, Bangladesh (icddr,b).

## Results

Of 1,923 patients hospitalized with acute jaundice, 148 (8%) had evidence of anti-HAV IgM antibodies in their serum, indicating an acute HAV infection (Table 1). Barisal reported the highest proportion of patients with HAV infection (20%); other hospitals had 3-7%. Males (9%) were more frequently affected compared to females (6%) (p=0.028). Younger patients aged 14 - 29 years were more likely to have HAV IgM antibodies (14%) compared to older age groups (0-3%) (p<0.001). Patients hospitalized with acute jaundice and HAV antibodies were also more likely to live in urban areas, have more education, and wealth than those without antibodies (Table 1).

**Table 1.**
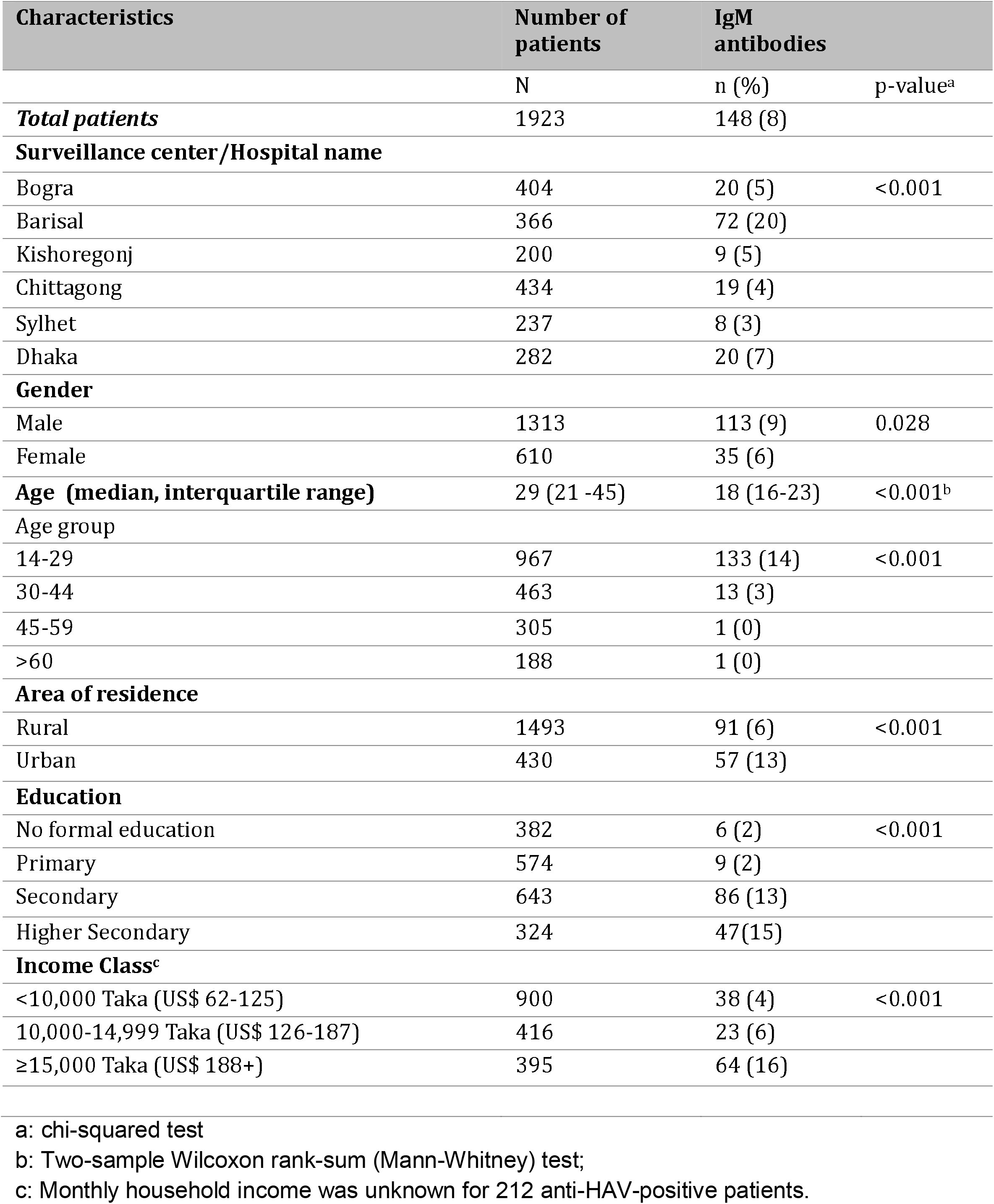
Hepatitis A IgM antibody test results by demographic characteristics of patients with acute jaundice in six tertiary hospitals in Bangladesh, December 2014– September 2017.

Patients with HAV IgM antibodies were hospitalized during every month of the study and from all hospital locations; no notable patterns were observed except for an apparent outbreak in Barisal during August - November 2016 when 36 patients with HAV infection were identified, representing 24% of all cases detected in this study (Figure 1).

**Figure 1:**
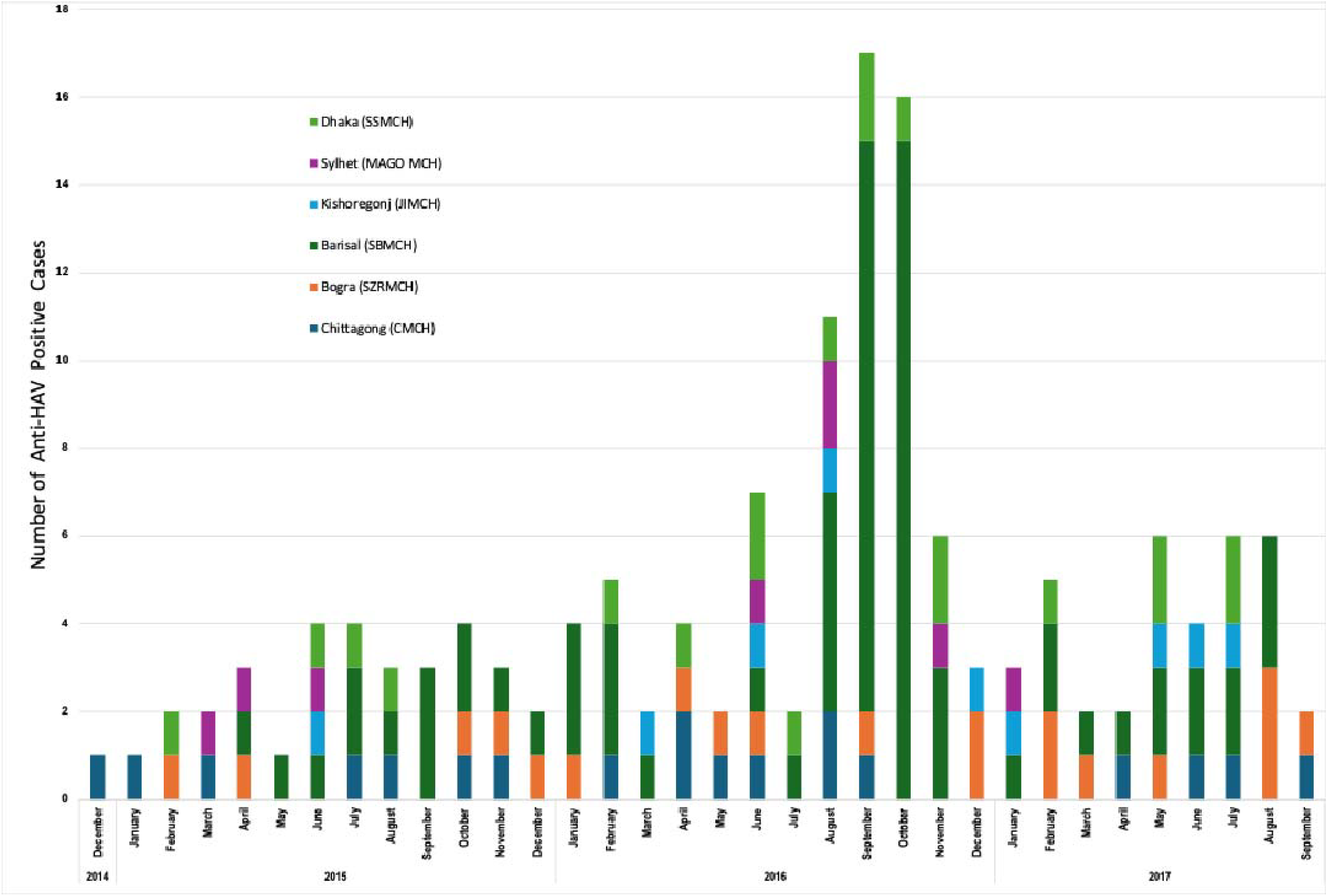
Stacked chart showing the number of patients with acute jaundice and IgM antibodies against hepatitis A virus admitted in six tertiary hospitals in Bangladesh, Dec 2014–September 2017.

All patients presented with yellow skin, yellow eyes, and dark-colored urine, per the surveillance case definition. Other symptoms, such as abdominal pain (45%), drowsiness (18%), altered mental status (2%), and unconsciousness (2%), were less common among patients with HAV compared to others (Table 2). Seventy-two percent of patients with HAV infection presented for care within 2 weeks of onset of illness, compared to 48% of other acute jaundice patients (Table 2). Six percent of patients with HAV infections also had anti-HEV IgM antibodies, and 20% HBsAg in serum, indicating co-infections. One patient had evidence of infection with all three viruses. Among the 305 deaths reported in patients with acute jaundice in this study, 1.3% (4 out of 305) had HAV infection (Table 2).

**Table 2.**
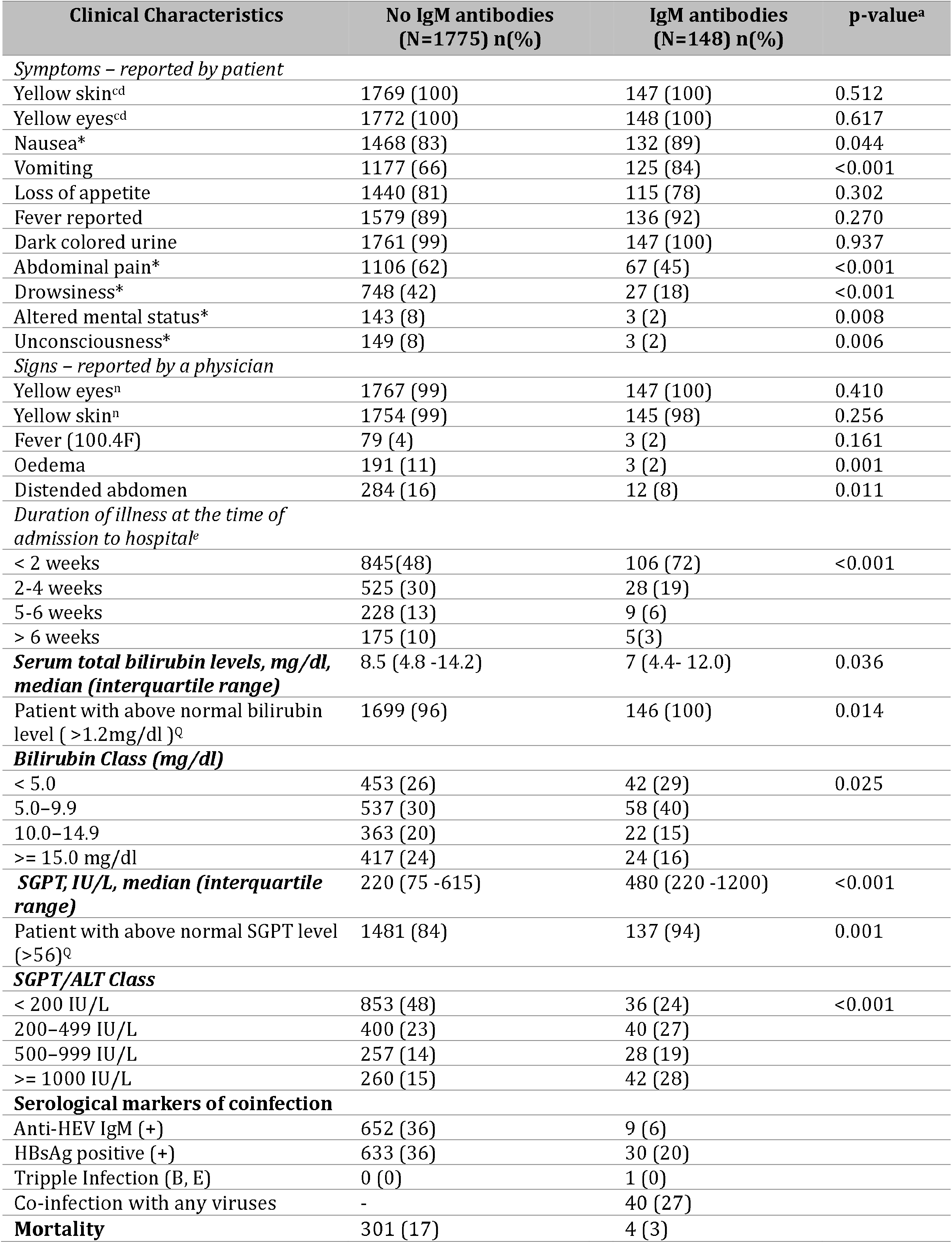
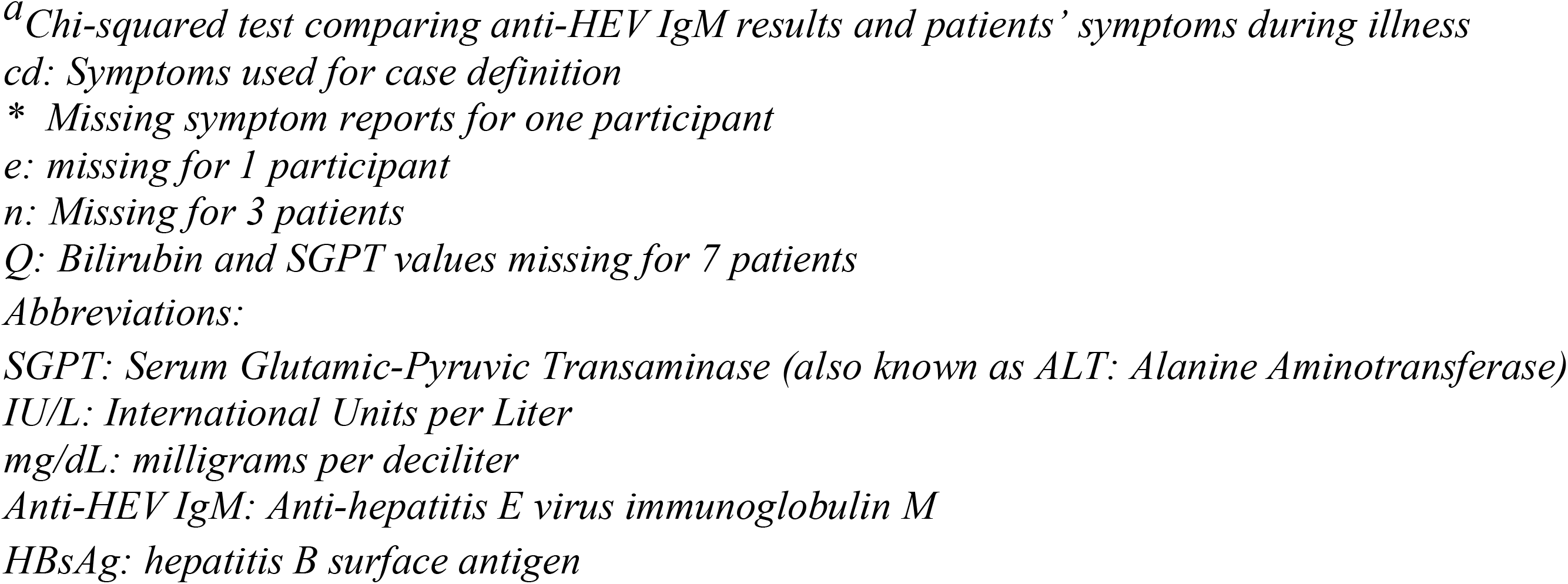
Signs and symptoms during illness among patients admitted in six tertiary hospitals in Bangladesh, December 2014– September 2017 by results from hepatitis A IgM antibody results.

| Clinical Characteristics | No IgM antibodies<br>(N=1775) n(%) | IgM antibodies<br>(N=148) n(%) | p-value<sup>a</sup> |
| --- | --- | --- | --- |
| *Symptoms – reported by patient* |  |  |  |
| Yellow skin<sup>cd</sup> | 1769 (100) | 147 (100) | 0.512 |
| Yellow eyes<sup>cd</sup> | 1772 (100) | 148 (100) | 0.617 |
| Nausea* | 1468 (83) | 132 (89) | 0.044 |
| Vomiting | 1177 (66) | 125 (84) | <0.001 |
| Loss of appetite | 1440 (81) | 115 (78) | 0.302 |
| Fever reported | 1579 (89) | 136 (92) | 0.270 |
| Dark colored urine | 1761 (99) | 147 (100) | 0.937 |
| Abdominal pain* | 1106 (62) | 67 (45) | <0.001 |
| Drowsiness* | 748 (42) | 27 (18) | <0.001 |
| Altered mental status* | 143 (8) | 3 (2) | 0.008 |
| Unconsciousness* | 149 (8) | 3 (2) | 0.006 |
| *Signs – reported by a physician* |  |  |  |
| Yellow eyes<sup>n</sup> | 1767 (99) | 147 (100) | 0.410 |
| Yellow skin<sup>n</sup> | 1754 (99) | 145 (98) | 0.256 |
| Fever (100.4F) | 79 (4) | 3 (2) | 0.161 |
| Oedema | 191 (11) | 3 (2) | 0.001 |
| Distended abdomen | 284 (16) | 12 (8) | 0.011 |
| *Duration of illness at the time of admission to hospital<sup>e</sup>* |  |  |  |
| < 2 weeks | 845 (48) | 106 (72) | <0.001 |
| 2-4 weeks | 525 (30) | 28 (19) |  |
| 5-6 weeks | 228 (13) | 9 (6) |  |
| > 6 weeks | 175 (10) | 5 (3) |  |
| **Serum total bilirubin levels, mg/dl, median (interquartile range)** | 8.5 (4.8 -14.2) | 7 (4.4- 12.0) | 0.036 |
| Patient with above normal bilirubin level (>1.2mg/dl)<sup>q</sup> | 1699 (96) | 146 (100) | 0.014 |
| **Bilirubin Class (mg/dl)** |  |  |  |
| < 5.0 | 453 (26) | 42 (29) | 0.025 |
| 5.0–9.9 | 537 (30) | 58 (40) |  |
| 10.0–14.9 | 363 (20) | 22 (15) |  |
| >= 15.0 mg/dl | 417 (24) | 24 (16) |  |
| **SGPT, IU/L, median (interquartile range)** | 220 (75 -615) | 480 (220 -1200) | <0.001 |
| Patient with above normal SGPT level (>56)<sup>q</sup> | 1481 (84) | 137 (94) | 0.001 |
| **SGPT/ALT Class** |  |  |  |
| < 200 IU/L | 853 (48) | 36 (24) | <0.001 |
| 200–499 IU/L | 400 (23) | 40 (27) |  |
| 500–999 IU/L | 257 (14) | 28 (19) |  |
| >= 1000 IU/L | 260 (15) | 42 (28) |  |
| **Serological markers of coinfection** |  |  |  |
| Anti-HEV IgM (+) | 652 (36) | 9 (6) |  |
| HBsAg positive (+) | 633 (36) | 30 (20) |  |
| Tripple Infection (B, E) | 0 (0) | 1 (0) |  |
| Co-infection with any viruses | - | 40 (27) |  |
| **Mortality** | 301 (17) | 4 (3) |  |

<sup>a</sup>Chi-squared test comparing anti-HEV IgM results and patients' symptoms during illness
cd: Symptoms used for case definition
\* Missing symptom reports for one participant
e: missing for 1 participant
n: Missing for 3 patients
Q: Bilirubin and SGPT values missing for 7 patients
SGPT: Serum Glutamic-Pyruvic Transaminase (also known as ALT: Alanine Aminotransferase)
IU/L: International Units per Liter
mg/dL: milligrams per deciliter
Anti-HEV IgM: Anti-hepatitis E virus immunoglobulin M

Four out of 148 patients with HAV infection died (3%); three of them had co-infections. Patients who died ranged in age from 22 - 70 years; two patients died in the hospital, the other two at home (Supplementary information).

## Discussion

This study of HAV infection among patients hospitalized with acute jaundice across six hospitals in Bangladesh between 2014-2017 found that 8% had IgM antibodies against HAV, which is lower than two other studies from Bangladesh during similar time periods (19% in 2014^6^ and 32% in 2012^8^). One possible reason for the lower prevalence in our study was the age of participants; our study focused on adults while the other studies enrolled all age groups and identified higher prevalence of HAV infection among children. While most studies do not investigate co-infections with other viral hepatitis viruses, our diagnostic approach identified that 27% of patients hospitalized with HAV infection were co-infected with either HEV, HBV, or both. Co-infections can increase the likelihood of severe infection and hospitalization^9^, and therefore, interventions to reduce HEV and HBV infections will also likely reduce hospitalizations associated with HAV infection. Importantly, 3 out of the 4 deaths among patients with HAV infection had co-infections, suggesting that preventing any of these infections could have prevented fatal outcomes. An analysis of this same surveillance dataset identified that among patients hospitalized with HEV infections, 9% (154/1765) had co-infections with either HAV or HBV ^7^. The majority of HEV coinfections were with HBV. Bangladesh introduced HBV vaccine into routine childhood immunizations during 2003-2005 and future generations might be at decreased risk for co-infections and the associated severity because of this intervention.^10^

Similar to other studies, we identified that younger patients and those from wealthier, more urban environments were more likely to have HAV infections. One explanation for this might be that early exposure to this pathogen continues to decline among these groups, likely due to improvements to living conditions and water systems. It is possible that people are reaching adulthood without being infected more commonly; however, earlier studies of adult hospitalizations from HAV infection are limited by sample size and geographic scope, making inferences about changes over time difficult. One study from Dhaka in 2004 – 2006 identified that 6% of adults seeking care for acute hepatitis had HAV infection, similar to our findings.^11^ A quarter of all patients with HAV infection in this study sought care during the second half of 2016 at the Barisal hospital surveillance site, suggesting an outbreak occurred there.

The burden of HAV in Bangladesh is dynamic and could be increasing, influenced by the prevalence of co-infections with other hepatitis viruses, and the age of infection. As birth cohorts immunized against HBV age into older adult ages, hospital admissions for HBV should decrease, and hospitalizations from co-infections with HEV and HAV may decrease as well. Studies to monitor changes in the causes of viral hepatitis hospitalizations over time could be useful for policy makers interested in reducing the burden of acute hepatitis.

## Data Availability

All data produced in the present study are available upon reasonable request to icddr,b.

## Acknowledgements

We are grateful to the collaborating hospitals for their interest, enthusiasm and efforts for this study. We gratefully acknowledge the contribution of the study participants and the surveillance medical officers. We are thankful to the field assistants of icddr,b and laboratory staff of IEDCR for their hard work in this study.

## Disclosures regarding conflicts of interest

The authors have no real or perceived conflicts of interest.

## Supplementary information

### Case descriptions of hepatitis A virus (HAV) infection related deaths

#### Patient 1

Patient 1, a pregnant female in her 20’s from a rural district of Sylhet, had a primary education and a household income of less than 10,000 Taka (US$ 125). She reported illness for ten days before she was hospitalized in June 2016. She reported symptoms such as yellow skin, yellow eyes, dark urine, abdominal pain, nausea, vomiting, and fever. Upon examination, the patient had yellow skin and yellow eyes. Laboratory tests revealed a bilirubin level of 4.6 mg/dL and an SGPT level of 56 IU/L, categorizing her bilirubin levels as above normal, while her SGPT levels were within normal limits. Testing indicated co-infection with HBV, evidenced by HBsAg. She was discharged seven days after hospitalization and died at home on the same day.

#### Patient 2

Patient 2 was a pregnant female in her 40’s from a rural district of Kishoreganj with a primary education and household income below 10,000 Taka (US$ 125). The patient reported illness for 15 days before hospitalization in March of 2016. She reported yellow skin, yellow eyes, dark urine, fever, convulsions, unconsciousness, and an altered mental status. Examination confirmed yellow skin and yellow eyes. Laboratory results showed a bilirubin level of 17.4 mg/dL and an SGPT level of 42 IU/L, categorizing her bilirubin levels as above normal, while SGPT levels were normal. She also had IgM antibodies against HEV, suggesting co-infection. She died after two days in hospital.

#### Patient 3

Patient 3, a female in her 70’s from the rural district of Kishoreganj with no formal education, had an illness duration of 60 days before hospitalization. Her household income was below 10,000 taka (US$125). The patient reported yellow skin, yellow eyes, dark urine, abdominal pain, nausea, and fever. The examination revealed yellow and yellow eyes. Her bilirubin level was 20.8 mg/dL, and SGPT was 159 IU/L, categorizing both levels as above normal. The patient tested negative for HEV IgM and HBsAg. She was admitted in June 2016 for nine days and died at home 39 days after being discharged.

#### Patient 4

Patient 4 was a male in his 30’s from a rural district of Joypurhat with secondary education and a household income below 10,000 Taka (US$125). His illness began 12 days prior to hospitalization. The patient reported yellow skin, yellow eyes, dark urine, nausea, and fever.

Examination confirmed yellow skin and yellow eyes. Bilirubin and SGPT levels were not measured. He tested positive for HEV IgM and negative for HBsAg, indicating a co-infection with HAV and HEV. He was admitted in May 2016 and died at home 10 days after he left the hospital. He did not exhibit unconsciousness or altered mental status, and details about the distended abdomen and diagnosis were not recorded.

